# Autoantibody screening in Guillain-Barré Syndrome

**DOI:** 10.1101/2021.05.10.21256964

**Authors:** Cinta Lleixà, Lorena Martín-Aguilar, Elba Pascual-Goñi, Teresa Franco, Marta Caballero, Jordi Diaz-Manera, Ricard Rojas-García, Noemí de Luna, Eduard Gallardo, Elena Cortés-Vicente, Joana Turón, Xavier Suárez-Calvet, Carlos Casasnovas, Christian Homedes, Gerardo Gutiérrez-Gutiérrez, María Concepción Jimeno-Montero, José Berciano, Maria José Sedano Tous, Tania Garcia-Sobrino, Julio Pardo-Fernandez, Celedonio Márquez-Infante, Iñigo Rojas-Marcos, Ivonne Jericó-Pascual, Eugenia Martínez-Hernández, Germán Morís de la Tassa, Cristina Domínguez-González, Laura Martínez-Martínez, Cándido Juárez, Isabel Illa, Luis Querol

## Abstract

Guillain-Barré Syndrome (GBS) is an acute inflammatory neuropathy with a heterogeneous presentation and pathogenesis. Serum antibodies against various gangliosides can be found in less than half of all patients in the acute phase of GBS but the target antigens remain unknown for the remaining half. Our work describes a comprehensive screening for serum autoantibodies targeting peripheral nerve tissue, cells, and purified antigens in a prospective GBS cohort including 100 patients. Our study confirms that (1) GBS patients display a very heterogeneous repertoire of autoantibodies targeting nerve cells and structures, (2) gangliosides are the most frequent antigens in GBS patients and have prognostic value, (3) a small subset of patients display antibodies targeting the myelin sheath, and (4) further antigen-discovery experiments are needed to elucidate other potential disease-specific autoantibodies in GBS.

## Introduction

Guillain-Barré Syndrome (GBS) is an acute inflammatory neuropathy with a heterogeneous presentation that includes diverse clinical variants.^1,2,3^ Diagnosis is based on clinical criteria; diagnostic biomarkers are not available for most patients.^4,5^ The exact immunopathogenic mechanisms of GBS are relatively unknown, but it is considered a paradigmatic post-infectious autoimmune disease. The response to intravenous immunoglobulins (IVIg) or plasma exchange support the role of autoantibodies in its pathogenesis.^6,7^

Anti-ganglioside antibodies are detected in half of GBS patients^8^. These autoantibodies arise via microbial molecular mimicry^9^ and the association of specific anti-ganglioside antibody reactivities and specific disease variants is well-established in the literature^10,11^, particularly the association of anti-GM1^12^ and GQ1b^13^ antibodies with the pure motor and Fisher syndrome variants of GBS respectively. In addition, the presence of antibodies targeting the GM1^12^ or GD1a^14,15^ gangliosides has also been associated with GBS prognosis. Antibodies against nodal and paranodal proteins (neurofascin 140/186^16^, neurofascin 155^17,18^, CASPR1^19,20^, and contactin 1^20^) have also been described in patients diagnosed of GBS. However, the target antigens remain unknown in a substantial proportion of GBS patients, particularly of the sensory-motor demyelinating variant, the most frequent in patients of European ancestry.

Considering the broad clinical and epidemiological spectrum of GBS, the diverse infectious triggers and the T-cell independent nature of the immune reaction leading to the appearance of autoantibodies^8^, we hypothesized that a broad repertoire of autoantibodies targeting diverse nerve components may be causing nerve pathology in GBS. This study aims to (1) screen for autoantibodies against known antigens, (2) screen for antibodies against human and rodent nerve cells and monkey nerve tissue; (3) describe the diversity of staining patterns and (4) perform clinical-immunological correlations, in a well-characterized GBS cohort.

## Results

### Baseline characteristics

We included 100 participants from 11 Spanish centres participating in the IGOS study. GBS patients had an average of 57.4 years and were predominantly men (57%). 65% of patients presented with the typical sensorimotor variant, 19% presented with a pure motor GBS variant, 10% with Miller Fisher syndrome (MFS), 5% with pure sensory variant and 1 patient with ataxic variant. Regarding nerve conduction studies, 59% of patients were classified as acute inflammatory demyelinating polyneuropathy (AIDP), 12% as acute motor axonal neuropathy (AMAN), 7% as acute motor-sensory axonal neuropathy (AMSAN), 8% as normal, and 14% as equivocal. Detailed epidemiological features of the cohort were described elsewhere.^21^

### Screening for known autoantibodies

None of the GBS patients included in the study reacted against the paranodal and nodal proteins tested (NF155, NF140, NF186, CNTN1 and CASPR1).

Sixty-one patients tested positive for, at least, one anti-ganglioside antibody (GM1, GM2, GM3, GD1b, GD3, aGM1, GT1a, GT1b and GQ1b). Of these, 40 had IgG antibodies, 3 had IgM antibodies, and 18 had antibodies from both isotypes. Detailed anti-ganglioside reactivities are shown in supplementary figure 1.

Most frequent anti-ganglioside antibodies in our cohort were aGM1, GM1, GD1b and GQ1b. Overall, IgG anti-aGM1 antibodies were detected in 40 % of patients; IgG and IgM anti-GM1 antibodies were detected in 27% and 15 % of patients respectively, IgG anti-GD1b antibodies in 30%, and IgG anti-GQ1b antibodies in 21 % patients.

### Antibodies targeting peripheral nerve neurons

ICC experiments with primary cultures of rat DRG neurons and human motor neurons derived from a neuroblastoma cell line were used to identify novel IgG and IgM reactivities against peripheral nerve neurons. The screening was performed in 100 serum samples from GBS patients and 90 serum samples from a control group (including healthy controls and patients with other neuromuscular diseases). ICC results were grouped in three separate categories: moderate to strong positives (including scores 2 and 3), all positives (including scores 1, 2 and 3), and negatives (score 0). Detailed results are shown in supplementary figure 2, figure 1 and table 1.

**Table 2.** Statistical analysis of DRG and neuroblastoma neurons ICC, and monkey peripheral nerve IHC.

|  | GBS patients (n = 100) |  | Controls (n=90 / 56) |  | p value |  |
| --- | --- | --- | --- | --- | --- | --- |
|  | Any reactivity | Strong reactivity | Any reactivity | Strong reactivity | Any reactivity | Strong reactivity |
| **Neuroblastoma neurons IgG** | 11 (11 %) | 2 (2 %) | 10/90 (11,1 %) | 0/90 (0,0 %) | >0,999 (ns) | 0,499 (ns) |
| **Neuroblastoma neurons IgM** | 28 (28 %) | 8 (8 %) | 11/90 (12,2 %) | 2/90 (2,2 %) | 0,011 (*) | 0,105 (ns) |
| **DRG neurons IgG** | 32 (32%) | 6 (6 %) | 6/90 (6,7 %) | 0/90 (0,0 %) | <0,0001 (***) | 0,030 (*) |
| **DRG neurons IgM** | 34 (34 %) | 11 (11 %) | 22/90 (24,4 %) | 2/90 (2,2 %) | 0,156 (ns) | 0,020 (*) |
| **Monkey peripheral nerve IgG** | 56 (56 %) | 17 (17 %) | 30/56 (53,6 %) | 3/56 (5,4 %) | 0,8669 (ns) | 0,0455 (*) |
| **Monkey peripheral nerve IgM** | 44 (44 %) | 10 (10 %) | 20/56 (35,7 %) | 6/56 (10,7 %) | 0,3964 (ns) | >0,9999 (ns) |

Comparison between GBS patients and controls. *Strong reactivity* includes scores 2 and 3, and *any reactivity* includes scores 1, 2 and 3. Fluorescence intensity scores were analysed using contingency analysis with the application of a Fisher's exact test, accepting an alpha-level of <0.05 to determine significance.

**Figure 1.**
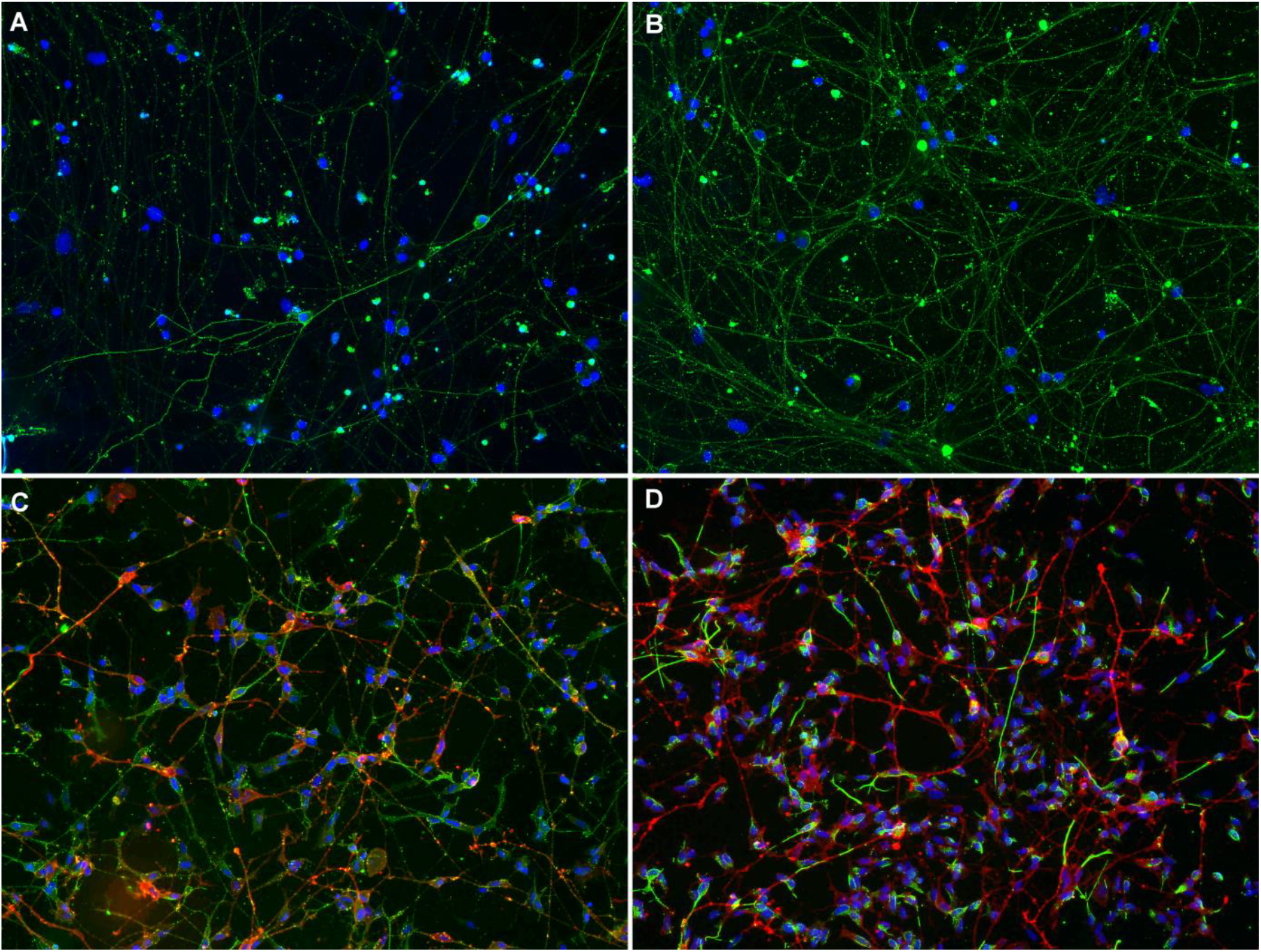
Reactivity against rat DRG neurons and human neuroblastoma-derived neurons. DRG neurons **(A-B)** stained with a GBS patient’s serum reacting moderately (score 2) in IgG **(A)**, and a GBS patient’s serum reacting strongly (score 3) in IgM **(B)**. Human neuroblastoma-derived neurons **(C-D)** stained in red with anti-panNeurofascin mAb, and in green with a GBS patient’s serum reacting moderately (score 2) in IgG **(C)** and a GBS patient’s serum reacting strongly (score 3) in IgM **(D)**.

Overall, 22 (22%) GBS patients reacted moderately or strongly against DRG or neuroblastoma neurons, whereas 4 (4.4%) controls reacted only moderately. These differences were statistically significant (p=0.0005).

Antibodies against DRG neurons appeared significantly more frequently in GBS patients than in controls (32% vs 6.7%, p<0.0001) taking all positive tests in account; the same happened if only moderate and strong positives were considered, both in IgG (6% vs 0%, p=0.03) and IgM experiments (11% vs 2.2%, p=0.02)

In neuroblastoma-derived neuron ICC experiments 28 (28%) samples from the GBS group showed IgM autoantibodies; of these 8 (8%) showed moderate or strong reactivity. These proportions were significantly higher than in the control group (12.2% and 2.2% respectively; p=0.011). Differences in autoantibody proportions between GBS patients and controls were not observed in neuroblastoma-derived neuron experiments when assessing IgG antibodies.

### Antibodies targeting peripheral nerve tissue

We analysed the full GBS cohort and 56 controls (a random subgroup).

We analysed the staining intensity of 6 different structures within the nerve, including nodes or paranodes, myelin from small myelinated fibers, myelin from large myelinated fibers, Schwann cells from unmyelinated fibers, large fiber axons, and small fiber axons. Staining patterns can be found in supplementary figure 3.

IgG and IgM reactivity against nerve tissue was frequently detected in GBS patients and controls. Overall, about 70% of GBS patients and controls sera bound to nerve structures. IgG and IgM from GBS patients reacted moderately in 17 (17%) and strongly in 10 (10%) against monkey nerve structures. In the control group IgG and IgM reacted moderately in 8 (14.3%) and strongly in 1 (1.8%) against monkey nerve structures. The difference between the amount of GBS patients and controls reacting moderately or strongly against monkey peripheral nerve was statistically significant (p=0.0455) only for IgG autoantibodies (Table 1).

Differences in IHC patterns of reactivity from GBS patients and controls were not statistically significant for any of the structures analysed (supplementary table 1). Nonetheless, some specific reactivity patterns were only detected in GBS patients and not in controls (figure 2). Eight (8%) GBS patients’ IgG reacted strongly against myelin, whereas only 2 controls showed weak reactivity against this structure. Moreover, we observed that 13 (13 %) GBS patients’ IgG had a strong reactivity against Schwann cells (myelinating and non-myelinating) while only one of the controls (1.8%) showed strong reactivity against Schwann cells (this difference is statistically significant; p=0.0192).

**Figure 2.**
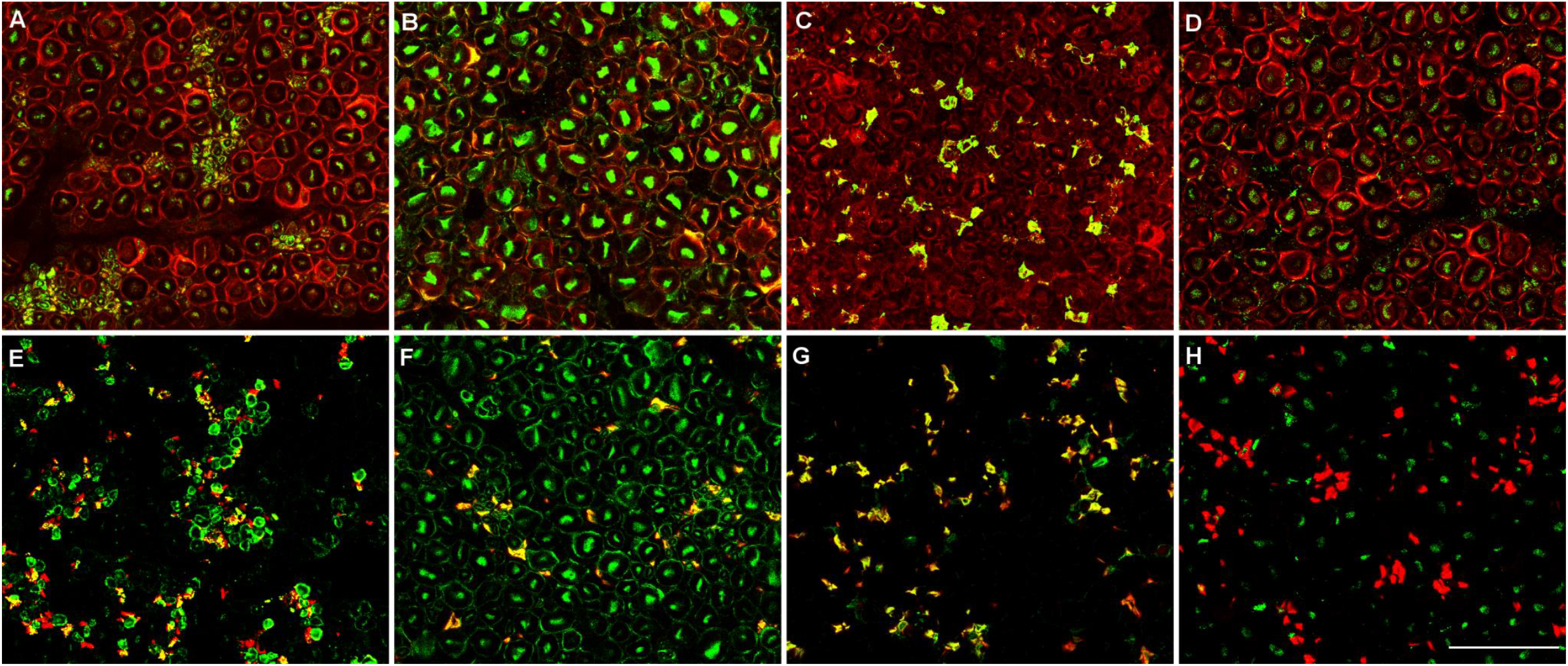
Reactivity against Schwann cells in peripheral nerve sections. Macaque peripheral nerve sections stained in red with S100 **(A-D)** or CD56 (NCAM) monoclonal antibody **(E-H)**, and in green with GBS patient’s sera reacting against myelin from small myelinated fibers **(A**,**E)**, myelin from large myelinated fibers **(B**,**F)**, and non-myelinating Schwann cells **(C**,**G). D** and **H** are stained with sera from negative controls.

### Combined autoantibody screening analysis

We also analysed if GBS patients with or without anti-ganglioside antibodies differed in the reactivity patterns in the peripheral nerve cell and tissue autoantibody screening experiments. No differences were found between those two groups (supplementary table 2), suggesting that the heterogeneity of the autoantibody repertoire appears even when a specific antigen is found.

We used a heatmap graph to represent all the autoantibody screening results performed in our GBS cohort (figure 3).^22^ This graph provides visual representation of the heterogeneity of the autoantibody repertoire in GBS sera.

**Figure 3.**
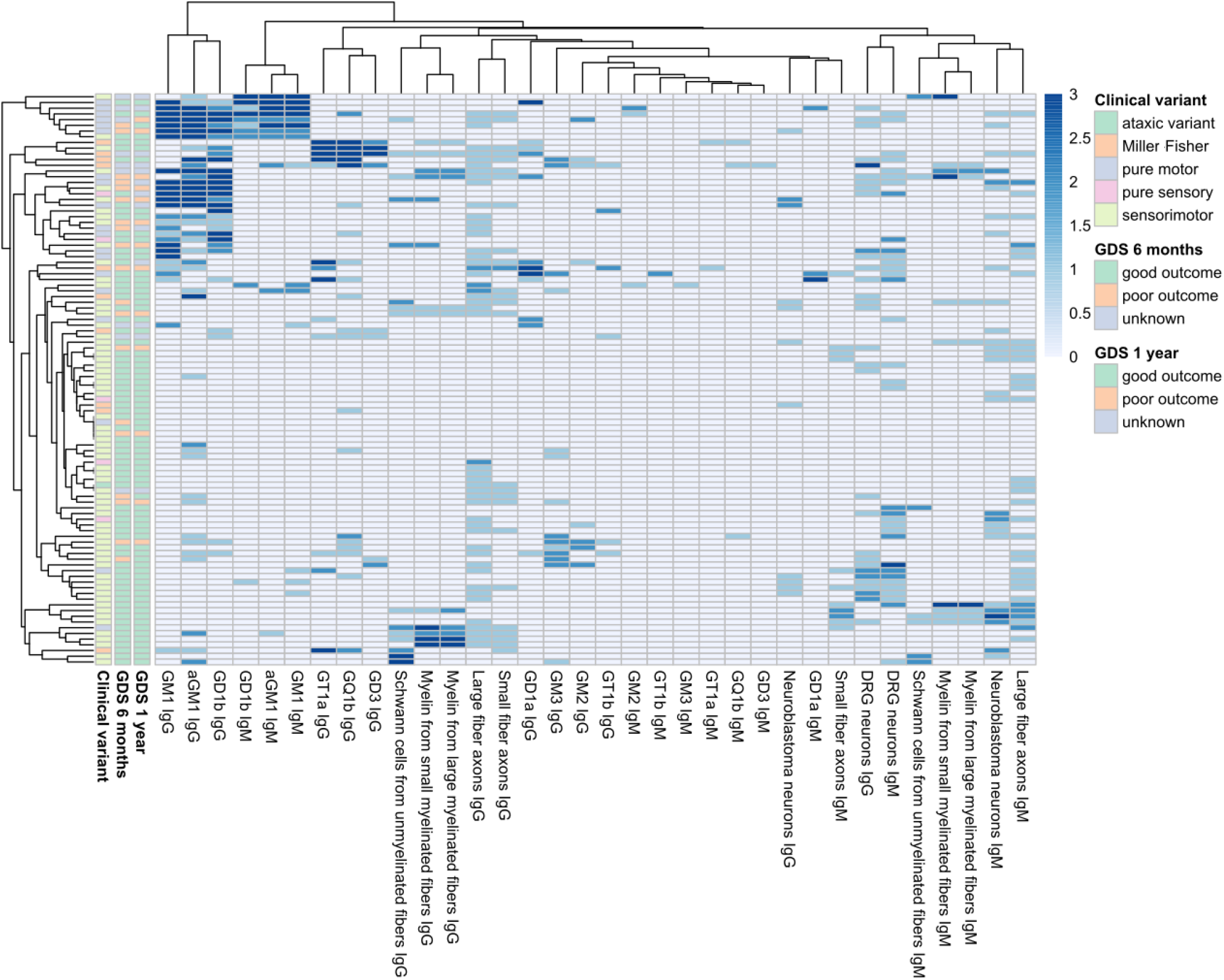
Heatmap showing all the screening performed in GBS patients. Patients and reactivities against anti-ganglioside antibodies, neuroblastoma-derived human motor neurons, murine dorsal-root ganglia neurons and monkey peripheral nerve tissue, are ordered according to Euclidean clustering. Each row represents one GBS patient. The score of the anti-ganglioside titre is indicated by the colour of the square (0=<1/1000, 1=1/1000 to 1/2500, 2=1/2500 to 1/12500, 3=>1/12500). The score of staining intensity in the other structures is indicated by the colour of the square (0=negative, 1=mild positive, 2=moderate positive, 3=strong positive). Columns in the left contain information related to the clinical variant and the outcome at 6 months and at 1 year of follow-up.

### Clinical correlations

Among patients with Miller Fisher syndrome, 8/10 (80%) had IgG anti-GQ1b antibodies, whereas in the rest of GBS patients only 14.4% (13/90) had these antibodies, usually in combination with other reactivities. IgG anti-GM1 antibodies were more frequently detected in patients with the pure motor variant than in those with other clinical variants (13/19 (68.4%) vs 14/81 (17.3%)); and in patients classified electrophysiologically as AMAN than in the rest of GBS patients (83.3% vs 19.3%). All these differences were statistically significant (p<0.0001).

When we analysed the general clinical characteristics of the subgroup of patients with strong IgG reactivity against Schwann cells (n=13), we did not observed any specific pattern that could distinguish them from the rest of the cohort. Ten (76,9%) of these patients presented with the sensorimotor clinical variant, whereas in the general cohort 65% presented this variant; and the proportions of nerve conduction studies subgroups were similar to those found in the general cohort (53.8% vs 59% of AIDP electrophysiological variant). Regarding the outcome, the percentage of patients having a good outcome at 6 months and 1 year are similar in the 2 groups (about 75%).

In the subgroup of patients with IgG or IgM reactivity against DRG neurons (n=14), we did not find clinical differences with the whole GBS cohort. Briefly, 71.4% of patients staining strongly DRG neurons were classified as AIDP, and 64.3% presented with the sensorimotor clinical variant.

We did not detect any difference in peripheral nerve cell and tissue reactivity patterns or frequencies between the samples collected before starting the treatment (62%) and those collected after the treatment (38%).

### Prognostic value of anti-ganglioside antibodies

First, we conducted a univariate analysis to select variables that were associated with the outcome. Patients with serum IgG anti-GM1 antibodies presented poorer outcomes than patients without the antibodies at 6 months (38.1% vs 16.1% (p=0.04)), and 1 year (35.3% vs 9.7% (p=0.014)). Anti-GD1a IgG antibodies were not associated with prognosis (table 2). For the multivariate analysis we included GM1 IgG, serum NfL levels, diarrhoea, age, and initial GDS.

**Table 2.**
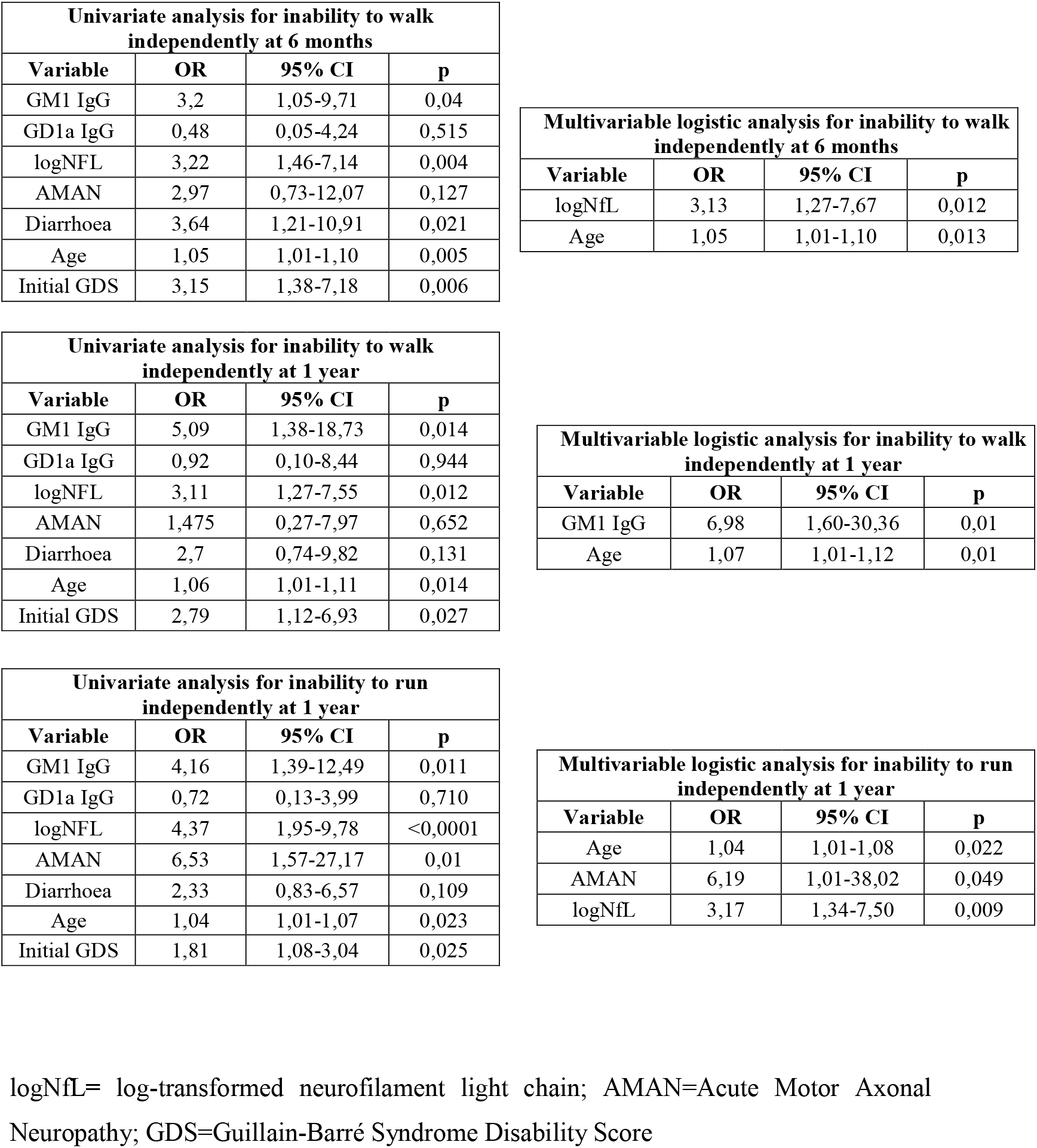
Association between baseline anti-GM1 and anti-GD1a antibodies and prognostic.

| Univariate analysis for inability to walk independently at 6 months |  |  |  |
| --- | --- | --- | --- |
| Variable | OR | 95% CI | p |
| GM1 IgG | 3,2 | 1,05-9,71 | 0,04 |
| GD1a IgG | 0,48 | 0,05-4,24 | 0,515 |
| logNFL | 3,22 | 1,46-7,14 | 0,004 |
| AMAN | 2,97 | 0,73-12,07 | 0,127 |
| Diarrhoea | 3,64 | 1,21-10,91 | 0,021 |
| Age | 1,05 | 1,01-1,10 | 0,005 |
| Initial GDS | 3,15 | 1,38-7,18 | 0,006 |

| Multivariable logistic analysis for inability to walk independently at 6 months |  |  |  |
| --- | --- | --- | --- |
| Variable | OR | 95% CI | p |
| logNfL | 3,13 | 1,27-7,67 | 0,012 |
| Age | 1,05 | 1,01-1,10 | 0,013 |

| Univariate analysis for inability to walk independently at 1 year |  |  |  |
| --- | --- | --- | --- |
| Variable | OR | 95% CI | p |
| GM1 IgG | 5,09 | 1,38-18,73 | 0,014 |
| GD1a IgG | 0,92 | 0,10-8,44 | 0,944 |
| logNFL | 3,11 | 1,27-7,55 | 0,012 |
| AMAN | 1,475 | 0,27-7,97 | 0,652 |
| Diarrhoea | 2,7 | 0,74-9,82 | 0,131 |
| Age | 1,06 | 1,01-1,11 | 0,014 |
| Initial GDS | 2,79 | 1,12-6,93 | 0,027 |

| Multivariable logistic analysis for inability to walk independently at 1 year |  |  |  |
| --- | --- | --- | --- |
| Variable | OR | 95% CI | p |
| GM1 IgG | 6,98 | 1,60-30,36 | 0,01 |
| Age | 1,07 | 1,01-1,12 | 0,01 |

| Univariate analysis for inability to run independently at 1 year |  |  |  |
| --- | --- | --- | --- |
| Variable | OR | 95% CI | p |
| GM1 IgG | 4,16 | 1,39-12,49 | 0,011 |
| GD1a IgG | 0,72 | 0,13-3,99 | 0,710 |
| logNFL | 4,37 | 1,95-9,78 | <0,0001 |
| AMAN | 6,53 | 1,57-27,17 | 0,01 |
| Diarrhoea | 2,33 | 0,83-6,57 | 0,109 |
| Age | 1,04 | 1,01-1,07 | 0,023 |
| Initial GDS | 1,81 | 1,08-3,04 | 0,025 |

| Multivariable logistic analysis for inability to run independently at 1 year |  |  |  |
| --- | --- | --- | --- |
| Variable | OR | 95% CI | p |
| Age | 1,04 | 1,01-1,08 | 0,022 |
| AMAN | 6,19 | 1,01-38,02 | 0,049 |
| logNfL | 3,17 | 1,34-7,50 | 0,009 |

logNfL= log-transformed neurofilament light chain; AMAN=Acute Motor Axonal Neuropathy; GDS=Guillain-Barré Syndrome Disability Score

We observed that having anti-GM1 IgG antibodies at baseline was independently associated with the inability to walk at 1 year of follow up, after a backward stepwise selection modelling (OR=6.98, 95% CI=1.6-30.36; p=0.01). However, the presence of anti-GM1 IgG antibodies was not independently associated with having a poor outcome at 6 months (table 2).

To analyse if anti-GM1 titres were associated with the GBS disability score, we performed a linear regression. We did not observe a positive correlation between antibody titres and disability at 6 months and at 1 year.

Finally, when we included the presence of anti-GM1 antibodies in our previously reported prognostic study^21^, we observed that having anti-GM1 IgG antibodies at baseline was associated with the inability to run at 1 year, but this association was not independent from the other known prognostic factors and sNfL, age and AMAN remained in the model as independent factors associated with residual disability at one year.

## Discussion

Our work describes a comprehensive autoantibody screening that provides experimental evidence of the heterogeneity of the autoantibody repertoire in patients fulfilling GBS diagnostic criteria. Our findings indicate that, other than gangliosides, single, common protein autoantigens are unlikely to be present in GBS patients.

Our study shows that GBS patients have a heterogeneous repertoire of autoantibodies targeting nerve cells and structures. Except for patients with anti-ganglioside antibodies and a minor subset of patients with antibodies targeting Schwann cells and the myelin sheath, this repertoire varies in frequency and intensity of staining, but it is not qualitatively different from controls. Antibodies targeting peripheral nerve cells of both IgG and IgM isotypes are significantly more frequent in patients than in controls, but no clear differences are seen when antibodies are tested using immunohistochemistry on monkey nerve preparations. Considering that whole nerve monkey preparations likely display antigens in a conformation that is phylogenetically closer to the one in human nerves, this may imply that autoantibodies targeting nerve structures are present at lower titers in normal human repertoire, that they arise as a natural epiphenomenon of a T-cell mediated damage and are not pathogenic, or that other autoantibodies, targeting different molecules (lipids, glycans) for which our techniques are not optimized, are yet to be discovered.

Whether these autoantibodies arise from a process of molecular mimicry, or from an unspecific and polyclonal activation of pre-existing B cells, remains unclear. The general absence of common patterns suggests the latter, but the well-established molecular mimicry process described in anti-ganglioside-associated GBS supports the former. In anti-GM1-associated GBS the sequence of pathogenic events includes an immune response to an infection leading to the appearance of antibodies cross-reacting with peripheral nerve and nerve root gangliosides and triggering post-infectious inflammation.^23^ Interestingly, in this screening we did not find clear differences in the reactivity patterns between GBS patients with or without anti-ganglioside antibodies, but we observed in both groups a higher amount of patients staining nerve structures than in controls. These findings suggest that the immune response is not restricted to the production of anti-ganglioside antibodies, but it is also generated against other peripheral nerve structures and supports the presence of a polyclonal, not antigen-driven, reactivation of a pre-existing repertoire that, in some patients, includes gangliosides or the concomitant activation (epitope spreading or bystander activation) of unspecific B-cells in addition to the ganglioside-driven specific response.

Previous studies in other inflammatory neuropathies such as chronic inflammatory demyelinating polyneuropathy (CIDP), showed that frequencies of reactivity against DRG neurons in CIDP patients did not differ from healthy controls^24^, in contrast with our results (shown in table 2). GBS and CIDP are clinically similar diseases so this difference supports the idea that a heterogeneous autoantibody response against multiple nerve antigens arises in GBS while this does not happen in chronic inflammatory and demyelinating neuropathies in which an specific, antigen-driven autoantibody response arises, as the recent discovery of the nodo-paranodal antibodies supports^25^.

Despite that, we observed that 13% of GBS patients showed strong IgG reactivity against Schwann cells of monkey peripheral nerve. This observation is in agreement with previous findings: Vallat et al detected that a significant percentage of CIDP and GBS patients (about 25%) presented with IgG or IgM reactivity against myelin and that the staining patterns on Schwann cells were diverse, suggesting that diverse myelin antigens are being recognized by autoantibodies.^26^

Our study also confirms, in a well-characterized GBS cohort, that gangliosides are the most frequent specific antigens in GBS patients and that they associate to specific disease variants. The value of testing anti-ganglioside antibodies in the GBS routine clinical care is controversial, but it is clear that some antibodies are associated with specific clinical phenotypes.^27^ IgG anti-GQ1b antibody is a diagnostic marker and a pathogenic antibody in MFS, and is often cross-reactive with GT1a^10^. Moreover, IgG anti-GM1 antibodies associate with the pure motor (clinical) and AMAN (electrophysiological) variants. Our results, with 80% of MFS patients having anti-GQ1b antibodies and 68.4% of pure motor patients having anti-GM1 antibodies at baseline, confirm these associations. However, despite being one of the largest cohorts in which antiganglioside antibodies have been tested, our study lacks power to find other potential associations previously described (anti-GD1b with acute ataxic neuropathy, anti-GT1a and pharyngo-cervico-brachial variant)^28,29^.

Some studies have reported a correlation between IgG anti-GM1 and anti-GD1a antibodies with a poor outcome in GBS patients.^30,14,15,23^ In our cohort, IgG anti-GD1a antibodies did not associate to a poor outcome of the disease^14^. However, our data confirms that IgG anti-GM1 antibody is an independent prognostic factor that associates with poor prognosis at 1 year, supporting that it may be a marker for long-term axonal damage. Whether the presence of complement-fixing anti-GM1 antibodies is the driver of this long-term disability, an important therapeutic question (that would enable the use of complement inhibitors in these patients), remains to be elucidated.

Although in this study we analysed the prognostic value of anti-ganglioside antibodies using the traditional outcome measures: inability to walk (GDS≥3) at 6 months and at 1 year, we have recently used the inability to run (GDS≥2). In this recent study we showed that high baseline sNfL were independently associated with inability to run at 1 year^21^. In agreement with these findings, we observed that including in the model the variable “presence of serum IgG anti-GM1 antibodies”, sNfL levels remained as an independent prognostic factor, whereas anti-GM1 antibodies did not. These results confirm that sNfL levels are a prognostic factor that informs better on axon status and, consequently, on long-lasting disability.

It is interesting to note that we did not find any patient with anti-nodal/paranodal antibodies (CNTN1, NF140, NF186, NF155 and CASPR1) in our GBS cohort. Although previous studies from other authors have found some GBS positive patients in their cohorts,^14-18^ and case-reports and series describe the association of anti-nodal/paranodal antibodies with aggressive inflammatory neuropathies frequently misdiagnosed as GBS, these antibodies are rare and we cannot rule out the possibility that they are present in other selected patients that our cohort failed to capture.

One of the limitations of our study is the number of patients and controls included. We have small groups of patients with similar staining patterns in which it is difficult to establish clear clinical-immunological correlations. Nevertheless, this is the first large prospective study assessing the autoantibody repertoire against peripheral nerve structures in GBS patients and antigen-identification experiments will follow in those patients with specific staining patterns that are absent in controls.

The existence of clear subgroups associated with anti-ganglioside antibodies, in contrast with the diversity in the new reactivities analysed, suggests that this apparent heterogeneity may be also due to technical caveats, because our study protocol is optimized for proteins and not for lipids or glycans. Moreover, other, not properly controlled factors, could have influenced heterogeneity in staining patterns (treatment, comorbidities…), and will need to be assessed in larger cohorts.

In conclusion, our study highlights the heterogeneity of the profile of autoantibodies targeting peripheral nerve structures, confirms gangliosides as the most frequent target antigens in the GBS autoantibody repertoire and their prognostic value in log-term GBS prognosis, and identifies small subsets of GBS patients with specific staining patterns in which further antigen-identification experiments could demonstrate novel and clinically relevant autoantibody reactivities in the future.

## Materials and methods

### Patients and controls

Serum samples from 100 GBS patients included in the Spanish cohort of the International GBS Outcome study (IGOS)^31^ were used in this screening. The IGOS is a multicentre, prospective, observational cohort study investigating factors that determine and predict the clinical course, subtype and outcome of GBS.^32^ Patients from the Spanish cohort were enrolled between February 2013 and January 2020. All patients fulfilled diagnostic criteria for GBS and were included within 2 weeks from onset of weakness. Serum samples were aliquoted and stored at -80°C until needed. In this study, we used serum samples extracted at baseline. Sixty-two (62%) of the baseline samples analysed were collected before starting treatment.

Clinical variants were defined as sensorimotor, pure motor, pure sensory, Miller Fisher syndrome (MFS) and ataxic. Nerve conduction studies results were classified as acute inflammatory demyelinating polyneuropathy (AIDP), acute motor axonal neuropathy (AMAN), acute motor-sensory axonal neuropathy (AMSAN), equivocal or normal. The outcome of all patients with GBS at 6 months and 1 year from disease onset were assessed using the GBS disability score (GDS), a widely accepted system for evaluating the functional ability of patients^33^. Patients unable to walk independently (≥3) at 6 months were defined as having a poor outcome in this study.

Additionally, serum samples from a control group (n=90) including 45 healthy controls and 45 patients with other neuromuscular disorders (23 ALS, 22 CMT) were included.

### Autoantibody screening protocol

Autoantibody screening experiments included antiganglioside antibody detection with ELISA, nodo/paranodal (NF155, NF140, NF186, CNTN1 and CNTN1/CASPR1 complex) antibody detection by ELISA and cell-based assays, immunocytochemistry using patient sera on neuroblastoma-derived human motor and murine dorsal-root ganglia neurons (IgG and IgM) and reactivity pattern assessment by immunohistochemistry on monkey sciatic nerve sections (IgG and IgM).

### Testing for nodo/paranodal antibodies

Autoantibodies against NF140, NF186, NF155, CNTN1 and CASPR1 were tested by ELISA.

Maxisorb 96-well ELISA plates (Thermo Fisher Scientific, NUNC, Denmark) were coated with 1μg/ml human recombinant CNTN1 protein (Sino Biological Inc., Georgia, USA), 1μg/ml NF155 protein (Origene, Maryland, USA), 1μg/ml NF140 protein (Sino Biological), 1 μg/ml NF186 protein (Origene) or 5μg/ml CASPR1 protein (R&Dsystems, MI, USA) overnight at 4 °C. Wells were blocked with 5% non-fat milk in PBS 0.1% Tween20 for 1 hour, incubated with sera diluted 1/100 in blocking buffer for 1 hour, and then incubated with peroxidase conjugated rabbit anti-human IgG secondary antibody (Invitrogen, CA, USA) for 1 hour at room temperature. ELISA was developed with tetramethylbenzidine solution (Biolegend, California, USA), and the reaction was stopped with 25% sulfuric acid. Optical density (OD) was measured at 450 nm in a Multiscan ELISA reader. Samples were considered positive by ELISA when they had a ΔOD higher than mean healthy control ΔOD plus two standard deviations.

Cell based assays were used as previously described^34^ as a second confirmatory technique for questionable cases. Briefly, mammalian expression vectors encoding human NF140, NF186, NF155, CNTN1 or CASPR1 were transfected into HEK293 cells using Lipofectamine 2000 (Invitrogen). Cells were then fixed with 4% paraformaldehyde and blocked. ICC experiments were performed using patient’s sera and appropriate primary and secondary antibodies.

### Testing for antiganglioside antibodies

Patients’ sera were screened for the presence of anti-ganglioside antibodies using a previously validated ELISA protocol^35^ as the general detection method, and thin layer chromatography^36^ for confirmatory experiments. Anti-ganglioside antibodies were considered positive at a 1/1000 titre.

### Rat dorsal root ganglia neurons immunocytochemistry

DRG were dissected from E16 rat embryos, dissociated and plated in glass-coverlips coated with laminin (Invitrogen) and poly-D-lysine (Sigma, MO, USA). Cells were grown in neurobasal medium (Gibco BRL, NY, USA) supplemented with B27 (Gibco), Glutamax (Gibco) and nerve growth factor (NGF) (Invitrogen). After 24 hours, cytosine arabinoside (ARA-C) (Sigma) and fluorouracil (5-FU) (Sigma) were added to the medium to remove fibroblasts. Then, medium was replaced every other day until reaching complete growth and differentiation of DRG neurons.

Live DRG neurons were incubated for 1 hour with patients’ sera diluted 1/100 (for IgG experiments) or 1/40 (for IgM experiments) in culture medium at 37°C. Cells were then fixed for 10 minutes with 4% PFA and incubated with secondary antibodies. Goat antihuman IgG (or IgM) AF488 (Molecular probes, Oregon, USA) were used as secondary antibodies at 1/1000 concentration.

Coverslips were mounted with Vectashield with DAPI and fluorescence signal intensity was scored in a 0–3 scale by two independent researchers. Images were obtained with an Olympus BX51 Fluorescence Microscope (Olympus Corporation, Tokyo, Japan). Animal procedures were performed according to a protocol approved by our Institution’s Animal Ethics’ Committee.

### Human neuroblastoma-derived neurons immunocytochemistry

SH-SY5Y cells were plated in glass-coverlips coated with laminin at 2.5 µg/ml (Invitrogen). Cells were grown in proliferation medium containing DMEM/F12 (1:1), fetal bovine serum (10%), L-glutamine (1%) and Sodium pyruvate (1%). After 24 hours, proliferation medium was replaced by differentiation medium containing Neurobasal (Gibco) supplemented with B27 (Gibco), Glutamax (Gibco), nerve growth factor (Invitrogen) and retinoic acid at 10 µM (Sigma). Then, medium was replaced every other day until full differentiation was achieved. On days 5 or 6 of differentiation, cells were fixed for 15 minutes with paraformaldehyde 4 %; and blocked with 5% normal goat serum in PBS; followed by incubation with patients’ sera at 1/40 (for IgM) or 1/100 (for IgG). To observe the correct differentiation of the cells we also incubated them with chicken anti-panNeurofascin mAb (R&Dsystems) at 1/200. Goat anti-chicken IgG AF594 and goat anti-human IgG AF488 or goat anti-human IgM AF488 (Molecular Probes) were used as secondary antibodies at 1/1000 concentration.

Coverslips were mounted with Vectashield with DAPI and fluorescence signal intensity was scored in a 0–3 scale by two independent researchers. Images were obtained with an Olympus BX51 Fluorescence Microscope.

### Peripheral nerve immunohistochemistry

Macaque peripheral nerve tissue slides (Inova Diagnostics, Inc., San Diego, CA) were blocked with 5% normal goat serum in PBS; followed by incubation with patients’ sera at 1:10 (for IgM) or 1:20 (for IgG). Monkey-adsorbed goat anti-human IgG AF488 (Southern Biotech, Alabama, US) or goat anti-human IgM AF488 (Molecular Probes) were used as secondary antibodies at 1/500 concentration. Finally, slides were mounted with Fluoromount medium (Sigma) and examined by two independent observers. Immunostaining patterns were analysed scoring fluorescence signal intensity of each nerve structure in a 0-3 scale. The nerve structures analysed were: nodes or paranodes, myelin from small myelinated fibers, myelin from large myelinated fibers, Schwann cells from unmyelinated fibers, large fiber axons, and small fiber axons. Reactivity against other non-nerve structures (fibroblasts, connective tissue, vessels) was not considered in the analysis.

To further study the staining patterns, peripheral nerve tissue slides were coated with mouse anti-human CD56 antibody (Becton Dickinson, New Jersey, USA) at 1:50 to stain non-myelinating Schwann cells (Remak bundles); or with rabbit anti-human S100 antibody (Abcam, Cambridge, UK) at 1:50 to stain myelinating Schwann cells. Goat anti-mouse IgG AF594 (for CD56), and goat anti-rabbit IgG AF594 (for S100) were used as secondary antibodies at 1/500 concentration.

Images were acquired using Leica TSC SP5 confocal microscope.

### Statistical analysis

Results were analysed by GraphPad Prism v8.0 (GraphPad Software). Statistical comparison of proportions among groups was performed using contingency analysis with the application of Chi-square and a two-tailed Fisher’s exact test, accepting an alpha-level <0.05 for statistical significance. To represent the results and perform clustering of our data we performed heatmap diagrams using Clustvis web tool.^22^

To investigate the association between anti-ganglioside antibodies and prognosis we used STATA and we performed a multivariable logistic regression analysis to predict the inability to walk at 6 months and at 1 year of follow-up (GDS≥3). First, we conducted a stepwise backward regression modelling to select variables independently associated with the outcome. The variables introduced in our initial multivariable models were selected based on known prognostic factors: age, initial GDS, diarrhoea, acute motor axonal neuropathy (AMAN), serum NfL levels (analyzed in a previous study with the same cohort)^21^, serum anti-GM1 IgG antibodies and serum anti-GD1a IgG antibodies^37,38,21,39^. To perform the multivariable analysis we excluded patients with MFS, because our aim was to predict GBS prognosis and MFS is considered a different disease, including different pathophysiology, clinical presentation (it does not present with weakness), treatment (often untreated) and outcome (considered self-limiting and benign). Moreover, we added the “presence of serum anti-GM1 IgG antibodies” variable to our previously reported prognostic study^21^, performing a multivariable logistic regression analysis to predict the inability to run at 1 year of follow-up (GDS≥2).

Odds-ratios (OR) for the logistic regression analysis were reported with 95% confidence intervals and p values.

### Standard protocol approvals, registrations and patient consents

The study was approved by the Ethics Committee of the Hospital de la Santa Creu i Sant Pau and the local institutional review boards of participating hospitals or universities. All patients gave written informed consent to participate in the study.

## Data Availability

Unpublished data will be shared in an anonymized manner on request from any qualified investigator for purposes of replicating procedures and results.

## Acknowledgements

The authors would like to acknowledge the Department of Medicine at the Universitat Autònoma de Barcelona, the IGOS consortium and Adela Gómez for their support. We also would like to thank all our patients for their patience and collaboration, and the animals that were sacrified to perform this study.

## Author contributions

CL acquired the data, performed the experiments, analyzed the data and drafted the manuscript; TF performed anti-ganglioside antibodies experiments; LMA, EPG, JDM, RRG, ECV, JT, CC, CH, GGG, MCJM, JB, MJST, TGS, JPF, CMI, IRM, IJP, EMH, GMT and CDG acquired samples and data and revised the manuscript for intellectual content; MC, NL, EG, XSC, LMM, CJ and II revised the manuscript for intellectual content; LQ designed and conceptualized the study, interpreted the data and revised the manuscript for intellectual content.

## Additional information: Supplementary data

**Supplementary figure 1.**
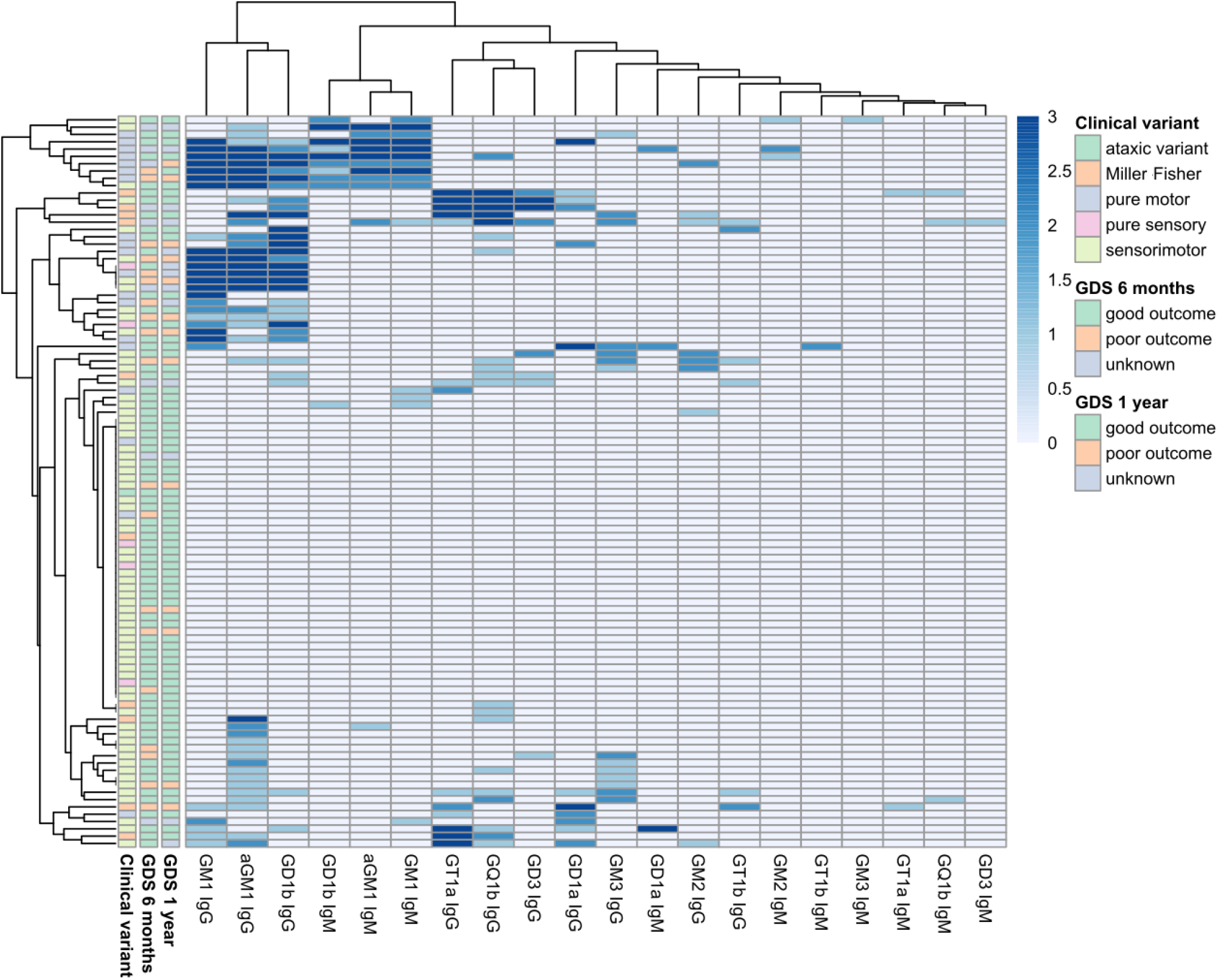
Heatmap showing anti-ganglioside antibodies in the GBS cohort. Patients and reactivities against anti-ganglioside antibodies are ordered according to Euclidean clustering. Each row represents one GBS patient. The score of the anti-ganglioside titre is indicated by the colour of the square (0 = <1/1000, 1 = 1/1000 to 1/2500, 2 = 1/2500 to 1/12500, 3 = >1/12500). Columns in the left contain information related to the clinical variant and the outcome at 6 months and at 1 year of follow-up.

**Supplementary figure 2.**
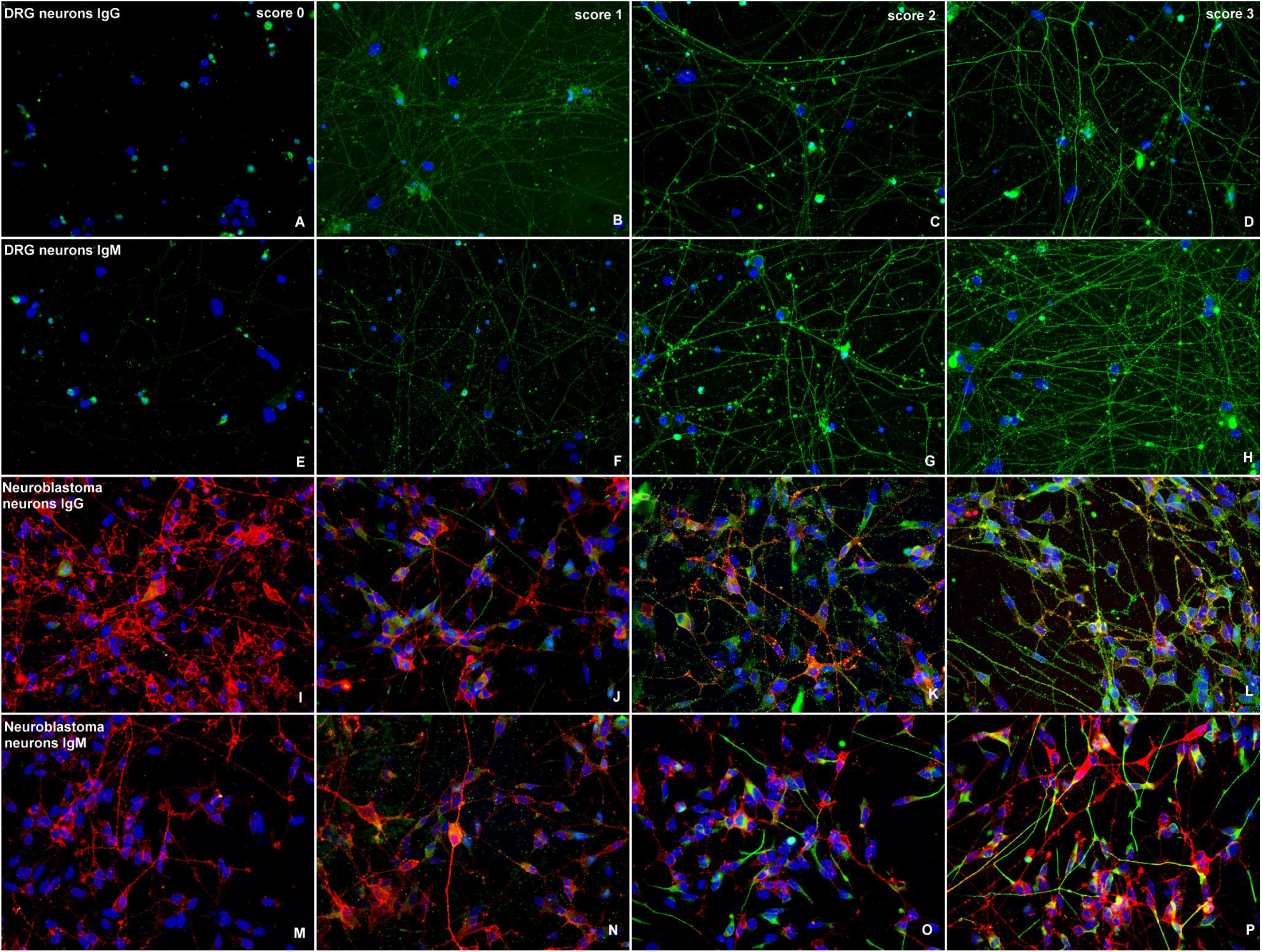
Staining paterns analized in ICC over rat DRG neurons and human neuroblastoma-derived neurons. DRG neurons (**A-H**) or neuroblastoma neurons (**I-P**) were stained with control or patient’s sera in green. Signal intensity of IgG (**A-D, I-L**) or IgM (**E-H, M-P**) reactivity was scored in a 0-3 scale (0=negative, 1=mild positive, 2=moderate positive, 3=strong positive). Neuroblastoma neurons were counterstained in red with anti-panNeurofascin mAb.

**Supplementary figure 3.**
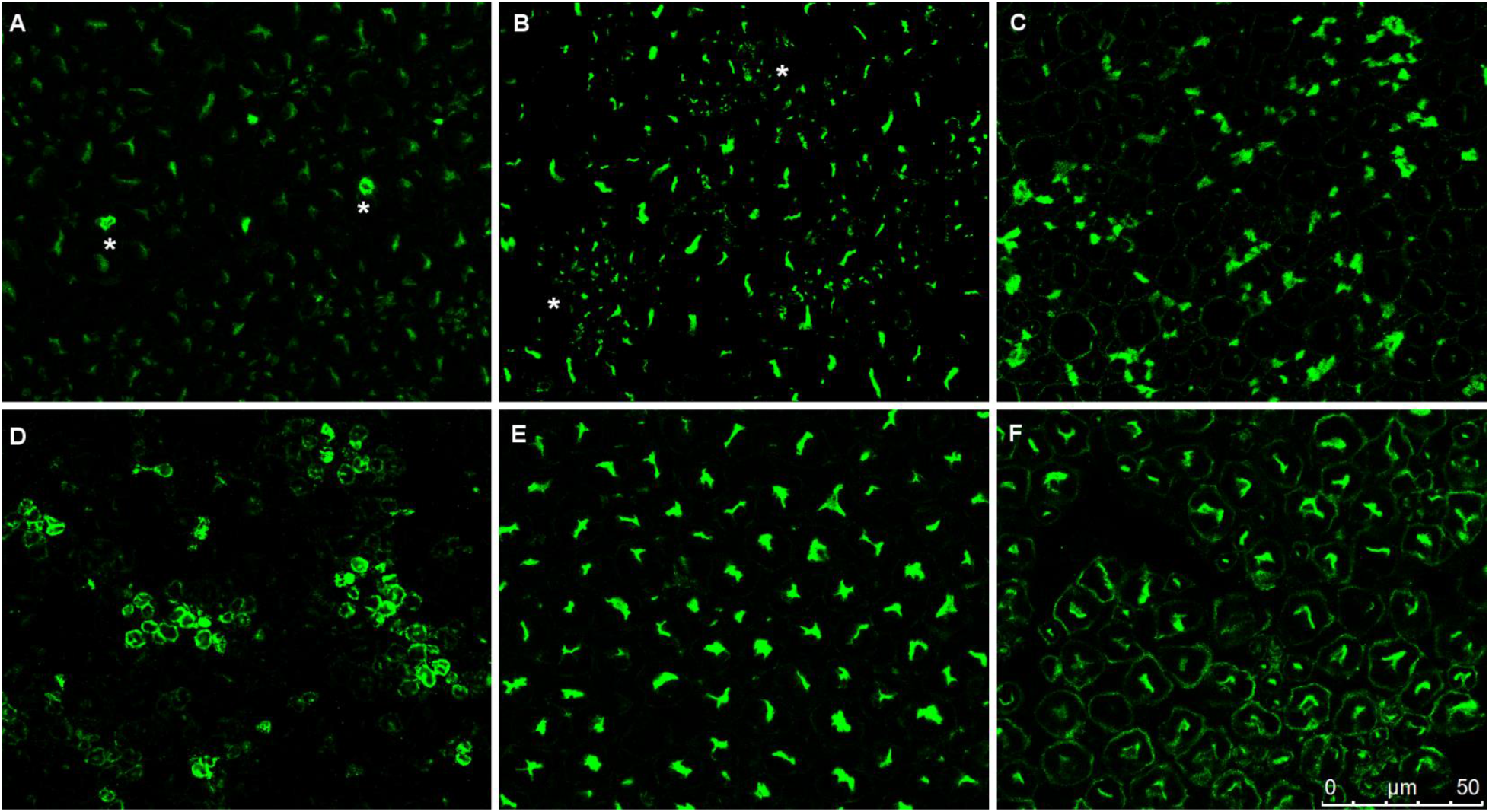
Staining paterns analized in IHC over monkey peripheral nerve. Macaque peripheral nerve transverse sections stained with CNTN1 positive CIDP patient’s serum reacting against paranodes **(A)**, small fiber axons **(B)**, non-myelinating Schwann cells **(C)**, myelin from small myelinated fibers**(D)**, large fiber axons **(E)**, and myelin from large myelinated fibers **(F)**.

**Supplementary table 1.**
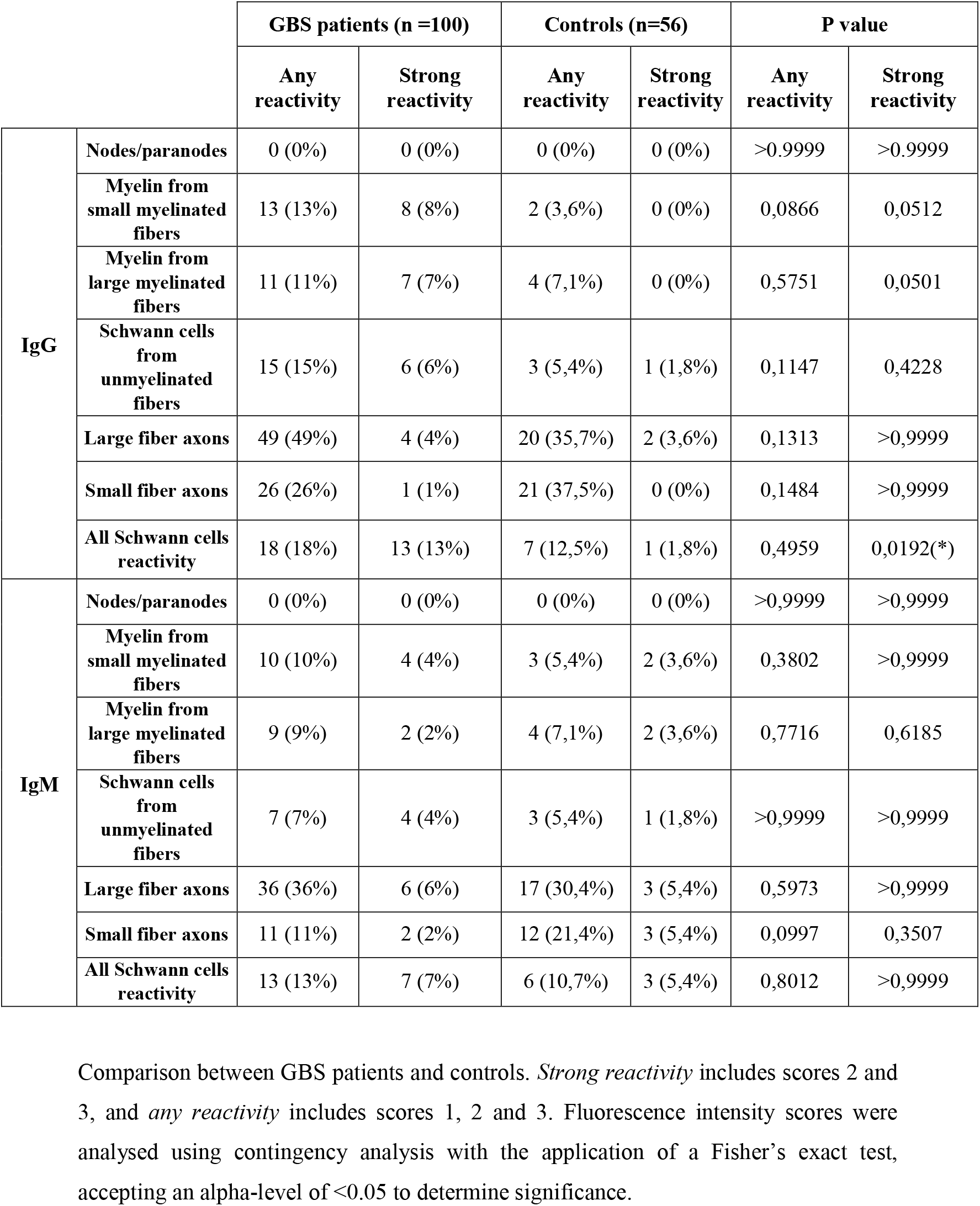
Statistical analysis of structures observed in IHC over monkey peripheral nerve.

**Supplementary table 2.** Statistical comparison between GBS patients with and without anti-ganglioside antibodies.

|  | GBS patients with anti-ganglioside antibodies (n = 61) |  | GBS patients without anti-ganglioside antibodies (n = 39) |  | P value |  |
| --- | --- | --- | --- | --- | --- | --- |
|  | Any reactivity | Strong reactivity | Any reactivity | Strong reactivity | Any reactivity | Strong reactivity |
| Neuroblastoma neurons IgG | 6 (9,8%) | 2 (3,3%) | 5 (12,8%) | 0 (0%) | 0,747 | 0,519 |
| Neuroblastoma neurons IgM | 13 (21,3%) | 3 (4,9%) | 15 (38,5%) | 5 (12,8%) | 0,072 | 0,256 |
| DRG neurons IgG | 22 (36,1%) | 5 (8,2%) | 9 (23,1%) | 1 (2,6%) | 0,191 | 0,400 |
| DRG neurons IgM | 24 (39,3%) | 8 (13,1%) | 10 (25,6%) | 3 (7,7%) | 0,196 | 0,521 |
| Monkey peripheral nerve IgG | 37 (60,7%) | 10 (16,4%) | 19 (48,7%) | 7 (17,9%) | 0,303 | >0,999 |
| Monkey peripheral nerve IgM | 22 (36,1%) | 6 (9,8%) | 22 (56,4%) | 6 (15,4%) | 0,063 | 0,530 |
| Nodes/paranodes IgG | 0 (0%) | 0 (0%) | 0 (0%) | 0 (0%) | - | - |
| Myelin from small myelinated fibers IgG | 7 (11,5%) | 5 (8,2%) | 6 (15,4%) | 3 (7,7%) | 0,561 | >0,999 |
| Myelin from large myelinated fibers IgG | 5 (8,2%) | 3 (4,9%) | 6 (15,4%) | 4 (10,3%) | 0,331 | 0,427 |
| Schwann cells from unmyelinated fibers IgG | 8 (13,1%) | 4 (6,6%) | 7 (17,9%) | 3 (7,7%) | 0,572 | >0,999 |
| Large fiber axons IgG | 32 (52,5%) | 3 (4,9%) | 17 (43,6%) | 1 (2,6%) | 0,418 | >0,999 |
| Small fiber axons IgG | 17 (27,9%) | 1 (1,6%) | 9 (23,1%) | 0 (0%) | 0,647 | >0,999 |
| All Schwann cells reactivity IgG | 10 (16,4%) | 7 (11,5%) | 8 (20,5%) | 6 (15,4%) | 0,605 | 0,561 |
| Nodes/paranodes IgM | 0 (0%) | 0 (0%) | 0 (0%) | 0 (0%) | - | - |
| Myelin from small myelinated fibers IgM | 4 (6,6%) | 3 (4,9%) | 6 (15,4%) | 1 (2,6%) | 0,182 | >0,999 |
| Myelin from large myelinated fibers IgM | 3 (4,9%) | 1 (1,6%) | 6 (15,4%) | 1 (2,6%) | 0,148 | >0,999 |
| Schwann cells from unmyelinated fibers IgM | 3 (4,9%) | 2 (3,3%) | 4 (10,3%) | 2 (5,1%) | 0,427 | 0,642 |
| Large fiber axons IgM | 17 (27,9%) | 2 (3,3%) | 19 (48,7%) | 4 (10,3%) | 0,054 | 0,205 |
| Small fiber axons IgM | 2 (3,3%) | 0 (0%) | 9 (23,1%) | 2 (5,1%) | 0,003 (**) | 0,150 |
| All Schwann cells reactivity IgM | 5 (8,2%) | 4 (6,6%) | 8 (20,5%) | 3 (7,7%) | 0,125 | >0,999 |

Comparison between GBS patients with and without anti-ganglioside antibodies. *Strong reactivity* includes scores 2 and 3, and *any reactivity* includes scores 1, 2 and 3. Fluorescence intensity scores were analysed using contingency analysis with the application of a Fisher's exact test, accepting an alpha-level of <0.05 to determine significance.

## References

1. Asbury, A. K. & Cornblath, D. R. Assessment of current diagnostic criteria for Guillain‐Barré syndrome. Ann. Neurol. 27, S21–S24 (1990).

2. Feasby, T. E. et al. An acute axonal form of guillain-barrée polyneuropathy. Brain 109, 1115–1126 (1986).

3. Wakerley, B. R., Uncini, A. & Yuki, N. Guillain-Barré and miller fisher syndromes - New diagnostic classification. Nat. Rev. Neurol. 10, 537–544 (2014).

4. Rinaldi, S. ’ syndrome Update on Guillain-Barr e. 112, 99–112 (2013).

5. Willison, H. J., Jacobs, B. C. & van Doorn, P. A. Guillain-Barré syndrome. Lancet (London, England) (2016) doi:10.1016/S0140-6736(16)00339-1.

6. Martín-Aguilar, L., Pascual-Goñi, E. & Querol, L. Autoantibodies in immune-mediated inflammatory neuropathies. Med. Clínica (English Ed. 153, 360–367 (2019).

7. Restrepo-Jiménez, P. et al. The immunotherapy of Guillain-Barré syndrome. Expert Opin. Biol. Ther. 18, 619–631 (2018).

8. Van Den Berg, B. et al. Guillain-Barré syndrome: Pathogenesis, diagnosis, treatment and prognosis. Nat. Rev. Neurol. 10, 469–482 (2014).

9. M. Green D. Advances in the management of Guillain-Barré Syndrome. Curr. Neurol. Neurosci. Rep. 2, 541–548 (2002).

10. Kaida, K. & Kusunoki, S. Antibodies to gangliosides and ganglioside complexes in Guillain-Barré syndrome and Fisher syndrome: Mini-review. J. Neuroimmunol. 223, 5–12 (2010).

11. Emilien, D. & Hugh, W. Diagnostic Utility of Auto Antibodies in Inflammatory Nerve Disorders. J. Neuromuscul. Dis. 2, 107–112 (2015).

12. Kuwabara, S. et al. IgG anti-GM1 antibody is associated with reversible conduction failure and axonal degeneration in Guillain-Barre syndrome. Ann. Neurol. 44, 202–208 (1998).

13. Willison, H. J., Veitch, J., Paterson, G. & Kennedy, P. G. E. Miller Fisher syndrome is associated with serum antibodies to GQ1b ganglioside. J. Neurol. Neurosurg. Psychiatry 56, 204–206 (1993).

14. Yamagishi, Y. et al. Serum IgG anti- - GD1a antibody and mEGOS predict outcome in Guillain- - Barré syndrome. 1–4 (2020) doi:10.1136/jnnp-2020-323960.

15. Rajabally, Y. A. & Uncini, A. Outcome and its predictors in Guillaine-Barré syndrome. J. Neurol. Neurosurg. Psychiatry 83, 711–718 (2012).

16. Devaux, J. J., Odaka, M. & Yuki, N. Nodal proteins are target antigens in Guillain-Barré syndrome. J. Peripher. Nerv. Syst. 17, 62–71 (2012).

17. Prüss, H., Schwab, J. M., Derst, C., Görtzen, A. & Veh, R. W. Neurofascin as target of autoantibodies in Guillain-Barré syndrome. Brain 134, 1–2 (2011).

18. Stengel, H. et al. Anti-pan-neurofascin IgG3 as a marker of fulminant autoimmune neuropathy. Neurol. Neuroimmunol. NeuroInflammation 6, 1–11 (2019).

19. Doppler, K. et al. Auto-antibodies to contactin-associated protein 1 (Caspr) in two patients with painful inflammatory neuropathy. Brain 139, 2617–2630 (2016).

20. Appeltshauser, L. Anti-paranodal antibodies and IgG subclasses in acute autoimmune neuropathy. Neurol. Neuroimmunol. Neuroinflammation (2020).

21. Martín-Aguilar, L. et al. Serum neurofilament light chain predicts long-term prognosis in Guillain-Barré syndrome patients. J. Neurol. Neurosurg. Psychiatry 1–8 (2020) doi:10.1136/jnnp-2020-323899.

22. Metsalu, T. & Vilo, J. ClustVis: A web tool for visualizing clustering of multivariate data using Principal Component Analysis and heatmap. Nucleic Acids Res. 43, W566–W570 (2015).

23. Van Koningsveld, R. et al. Infections and course of disease in mild forms of Guillain-Barré syndrome. Neurology 58, 610–614 (2002).

24. Querol, L. et al. Antibodies against peripheral nerve antigens in chronic inflammatory demyelinating polyradiculoneuropathy. Sci. Rep. 7, 1–9 (2017).

25. Querol, L., Devaux, J., Rojas-Garcia, R. & Illa, I. Autoantibodies in chronic inflammatory neuropathies: Diagnostic and therapeutic implications. Nat. Rev. Neurol. 13, 533–547 (2017).

26. Vallat, J. M. et al. Antibody-and macrophage-mediated segmental demyelination in chronic inflammatory demyelinating polyneuropathy: clinical, electrophysiological, immunological and pathological correlates. Eur. J. Neurol. 27, 692–701 (2020).

27. Leonhard, S. E. et al. Diagnosis and management of Guillain–Barré syndrome in ten steps. Nat. Rev. Neurol. 15, 671–683 (2019).

28. Pan, C. L., Yuki, N., Koga, M., Chiang, M. C. & Hsieh, S. T. Acute sensory ataxic neuropathy associated with monospecific anti-GD1B IgG antibody. Neurology 57, 1316–1318 (2001).

29. Koga, M., Yuki, N., Ariga, T., Morimatsu, M. & Hirata, K. Is IgG anti-GT1a antibody associated with pharyngeal-cervical-brachial weakness or oropharyngeal palsy in Guillain-Barre syndrome? J. Neuroimmunol. 86, 74–79 (1998).

30. Kuwabara, S. et al. Intravenous immunoglobulin therapy for Guillain-Barré syndrome ith IgG anti-GM1 antibody. Muscle Nerve 54–58 (2001).

31. Jacobs, B. C. et al. International Guillain-Barré Syndrome Outcome Study: protocol of a prospective observational cohort study on clinical and biological predictors of disease course and outcome in Guillain-Barré syndrome. J. Peripher. Nerv. Syst. 22, 68–76 (2017).

32. Doets, A. Y. et al. Regional variation of Guillain-Barré syndrome. Brain 141, 2866–2877 (2018).

33. Hughes, R. A. C., Newsom-Davis, J., Perkin, G. & Pierce, J. Controlled trial of prednisolone in acute polyneuropathy. Lancet (London, England) 750–753 (1978).

34. Siles, A. M. et al. Antibodies against cell adhesion molecules and neural structures in paraneoplastic neuropathies. Ann. Clin. Transl. Neurol. 5, 559–569 (2018).

35. Willison, H. J. et al. Inter-laboratory validation of an ELISA for the determination of serum anti-ganglioside antibodies. European Journal of Neurology vol. 6 71–77 (1999).

36. Ohanlon, G. M. et al. Peripheral neuropathy associated with anti-GM2 ganglioside antibodies: Clinical and immunopathological studies. Autoimmunity 32, 133–144 (2000).

37. Hiraga, A. et al. Recovery patterns and long term prognosis for axonal Guillain-Barré syndrome. J. Neurol. Neurosurg. Psychiatry 76, 719–722 (2005).

38. Kuwabara, S., Mori, M., Ogawara, K., Hattori, T. & Yuki, N. Indicators of rapid clinical recovery in Guillain-Barré syndrome. J. Neurol. Neurosurg. Psychiatry 70, 560–562 (2001).

39. Hadden, R. D. M. et al. Preceding infections, immune factors, and outcome in Guillain-Barré syndrome. Neurology 56, 758–765 (2001).

